# Quantifying individual nociceptive sensitivity to optimise analgesic trials in infants

**DOI:** 10.1101/2020.12.08.20246058

**Authors:** Maria M. Cobo, Caroline Hartley, Deniz Gursul, Foteini Andritsou, Marianne van der Vaart, Gabriela Schmidt Mellado, Luke Baxter, Eugene P. Duff, Miranda Buckle, Ria Evans Fry, Gabrielle Green, Amy Hoskin, Richard Rogers, Eleri Adams, Fiona Moultrie, Rebeccah Slater

**Affiliations:** Department of Paediatrics, University of Oxford, Oxford OX3 9DU, U.K.; Universidad San Francisco de Quito USFQ, Colegio de Ciencias Biologicas y Ambientales, Quito 170901, Ecuador; Wellcome Centre for Integrative Neuroimaging, University of Oxford OX3 9DU, UK; Newborn Care Unit, John Radcliffe Hospital, Oxford University Hospitals NHS Foundation Trust, Oxford OX3 9DU, UK; Department of Anaesthetics, John Radcliffe Hospital, Oxford University Hospitals NHS Foundation Trust, Oxford OX3 9DU, UK

## Abstract

Despite the high burden of pain experienced by hospitalised infants there are few analgesics with proven efficacy. Testing analgesics in infants is experimentally and ethically challenging and minimising the number of infants required to demonstrate efficacy is essential. EEG-derived measures of noxious-evoked brain activity can be used to assess analgesic efficacy, however, as variability exists in infant’s responses to painful procedures, large sample sizes are often required. Here we present a novel experimental paradigm to account for individual differences in baseline nociceptive sensitivity which can be used to improve the design of analgesic trials in infants. The paradigm is developed and tested across four studies using clinical, experimental and simulated data (99 infants). We provide evidence of the efficacy of both gentle brushing and paracetamol, substantiating the need for randomised controlled trials of these interventions. This work provides an important step towards safe, cost-effective clinical trials of analgesics in infants.

## Introduction

Considering the short-term stress and long-term neurodevelopmental impact associated with repeated pain exposure in early life (Brummelte et al., 2012; Chau et al., 2019; Morison et al., 2001; Vinall et al., 2014), effective pain relief is crucial in neonatal intensive care (Hall and Anand, 2014; Lim and Godambe, 2017). Nevertheless, as a result of the implicit challenges of measuring pain in infants, and the ethical and experimental challenges of conducting neonatal clinical trials, few analgesics have proven efficacy in this population (Allegaert, 2017; Moultrie et al., 2017; Slater et al., 2020). Participants in clinical trials risk exposure to potential adverse effects, and therefore every effort should be made to minimise the sample size necessary to demonstrate efficacy (EMA, 2001). However, as factors such as age (Fabrizi et al., 2011; Green et al., 2019; Hartley et al., 2016), prior pain experience (Ozawa et al., 2011; Slater et al., 2010a), stress (Jones et al., 2017), sex (Bartocci et al., 2006; Verriotis et al., 2018), illness (Ranger et al., 2011) and behavioural state (Slater et al., 2006) influence pain sensitivity, large sample sizes are often required to account for between-subject variability (Anand et al., 2004; Ancora et al., 2013; Hartley et al., 2018; Kabataş et al., 2016; Sindhur et al., 2020; Taddio et al., 2006).

In adult studies, cross-over trial designs are often used to minimise sample sizes by reducing between-subject variability (Cooper et al., 2016). However, this approach may not be appropriate when studying pain in infants as painful medical procedures can only be performed when clinically necessary and within-subject variables that influence pain can change dramatically across sequential test occasions. One approach used to balance demographic characteristics or other prognostic factors across treatment groups in clinical trials is to stratify infants across treatment arms, and to adjust for these factors in the statistical analysis (McEntegart, 2003). While this can improve comparability across groups for recognised factors, many unknown variables likely influence pain sensitivity and a more nuanced approach to account for individual differences in nociceptive sensitivity could be more effective in reducing sample sizes. In analgesic studies performed in adults, individual pain thresholds can be identified by applying graded increments of experimental stimulus intensity until pain is reported by the participants. This can be used to stratify treatment groups (Demant et al., 2014; Smith et al., 2017; Vollert et al., 2017) or statistically correct for variability in baseline pain thresholds (Lane et al., 2010; Sanga et al., 2013). In infants, application of graded non-noxious stimuli such as von Frey hairs have previously been used to identify limb reflex withdrawal thresholds (Andrews and Fitzgerald, 2002, 1999, 1994; Kuhne et al., 2012) but these have not been used as a baseline measure in analgesics clinical trials.

In the absence of a validated objective biomarker of pain (Davis et al., 2020), EEG-derived measures of brain activity may provide a valuable surrogate marker of pain by measuring the noxious-evoked activation of the cortex. In adults, brain activity during painful procedures is strongly correlated with verbal reports (Coghill et al., 2017); and in infants a template of noxious-evoked brain activity that discriminates between noxious and non-noxious procedures has been previously characterised and validated (Hartley et al., 2017). Here we develop and test an experimental paradigm that assesses individual nociceptive sensitivity in infants by measuring noxious-evoked brain activity in response to a low-intensity experimental noxious stimulus, and demonstrate that accounting for this measure of baseline nociceptive sensitivity substantially reduces the sample sizes required in neonatal studies of analgesic efficacy. In Study 1, we demonstrate that the magnitude of noxious-evoked brain activity in response to an experimental stimulus correlates with the magnitude of brain activity evoked by the clinical procedure and thus reflects baseline nociceptive sensitivity. In Study 2, we use simulated data to demonstrate the increased statistical power that can be achieved by including baseline nociceptive sensitivity as a covariate when analysing the effect of an intervention in small samples. In Study 3, we test this novel paradigm using a non-pharmacological pain-relieving intervention of known efficacy - gentle touch - prior to heel lancing. Finally, in Study 4 we investigated the analgesic efficacy of oral paracetamol given prior to immunisation in prematurely-born infants. Overall, we demonstrate that measuring and accounting for baseline nociceptive sensitivity could improve the design of analgesic efficacy investigations for this patient population.

## Results

### Study 1: Characterising individual baseline nociceptive sensitivity using brain activity in infants

We hypothesised that a measure of infant baseline nociceptive sensitivity could be used to account for inter-individual variability in noxious-evoked brain activity in studies of analgesic efficacy (nociceptive sensitivity paradigm, Figure 1) and predicted that this would reduce the sample sizes needed in clinical trials. In order to use noxious-evoked brain activity in response to a mild experimental noxious stimulus as a continuous measure of baseline nociceptive sensitivity, it must be significantly and strongly correlated with the response to the clinically-required procedure. In Study 1, we therefore assessed the feasibility and initial validity of the paradigm by investigating the relationship between individual responses of term infants to experimental noxious stimuli and a subsequent clinically-required heel lance. This was a retrospective study presenting previously unpublished data from term infants studied between 2014 and 2015 who had received both a heel lance and experimental noxious stimuli on the same test occasion (n=9). Whilst this sample size is small this was a feasibility study and this relationship is retested in Study 3. The magnitudes of the noxious-evoked brain activity evoked by stimulating an infant’s foot with a controlled mild experimental noxious stimulus (force = 64 mN; magnitude range: 0.15 - 0.62, Figure 2A) was strongly correlated with the magnitude of the noxious-evoked brain activity generated by a clinically-required heel lance (range: −0.07 – 2.34) in the same infants (p=0.0025, R^2^=0.77, Figure 2A). Therefore, application of a mild experimental noxious stimulus prior to performing a clinically-required painful procedure could provide a novel measure of infant baseline nociceptive sensitivity, which could be used as a covariate in studies of analgesic efficacy to account for inter-individual variability in pain responses (Figure 1). In the following sections, we simulate and test the impact of applying this novel paradigm in studies investigating the efficacy of pain-relieving interventions.

**Fig. 1.**
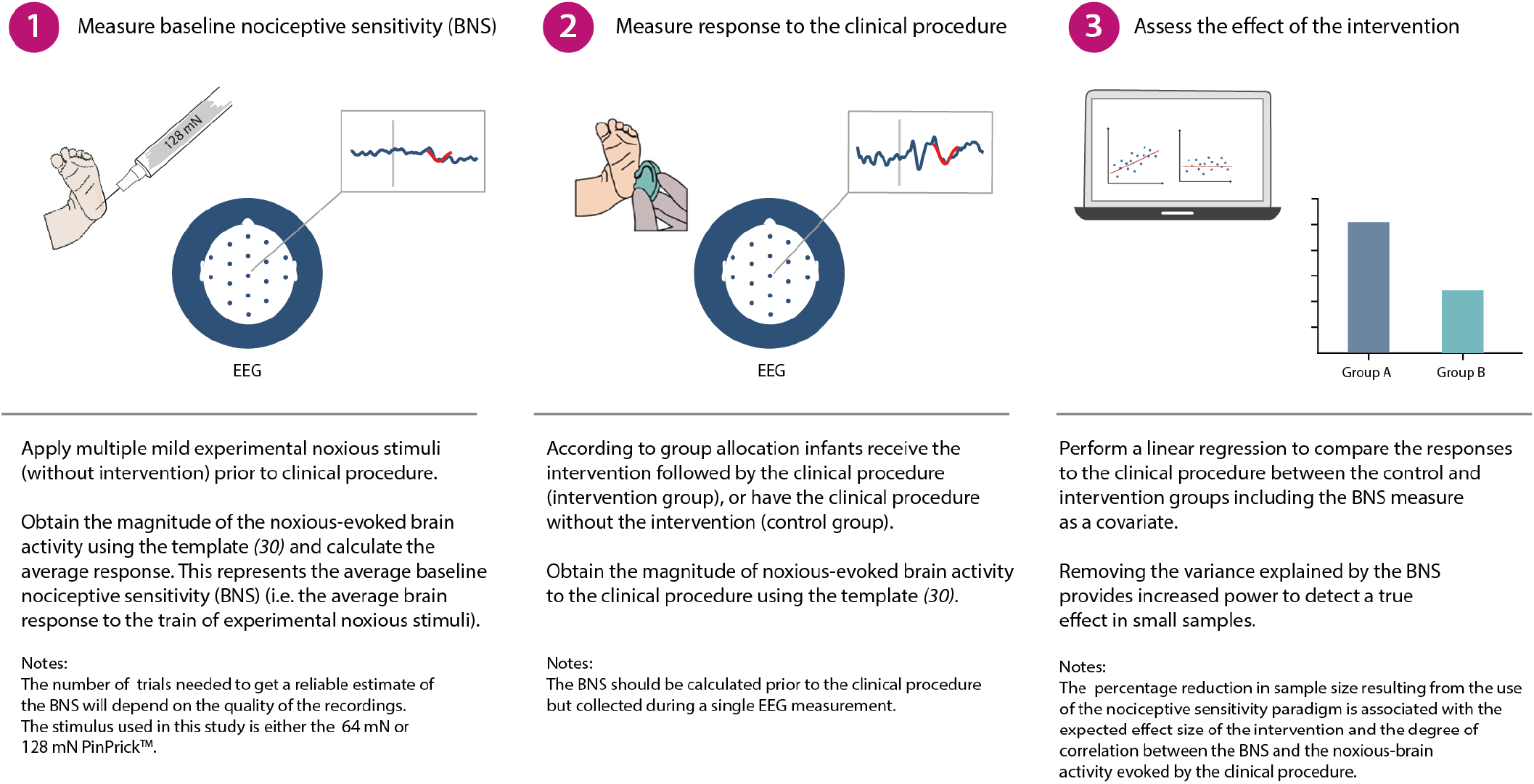
Nociceptive sensitivity paradigm explained. Schematic representation of the nociceptive sensitivity paradigm components. A brief description of each step is included with additional explanatory notes.

**Fig. 2.**
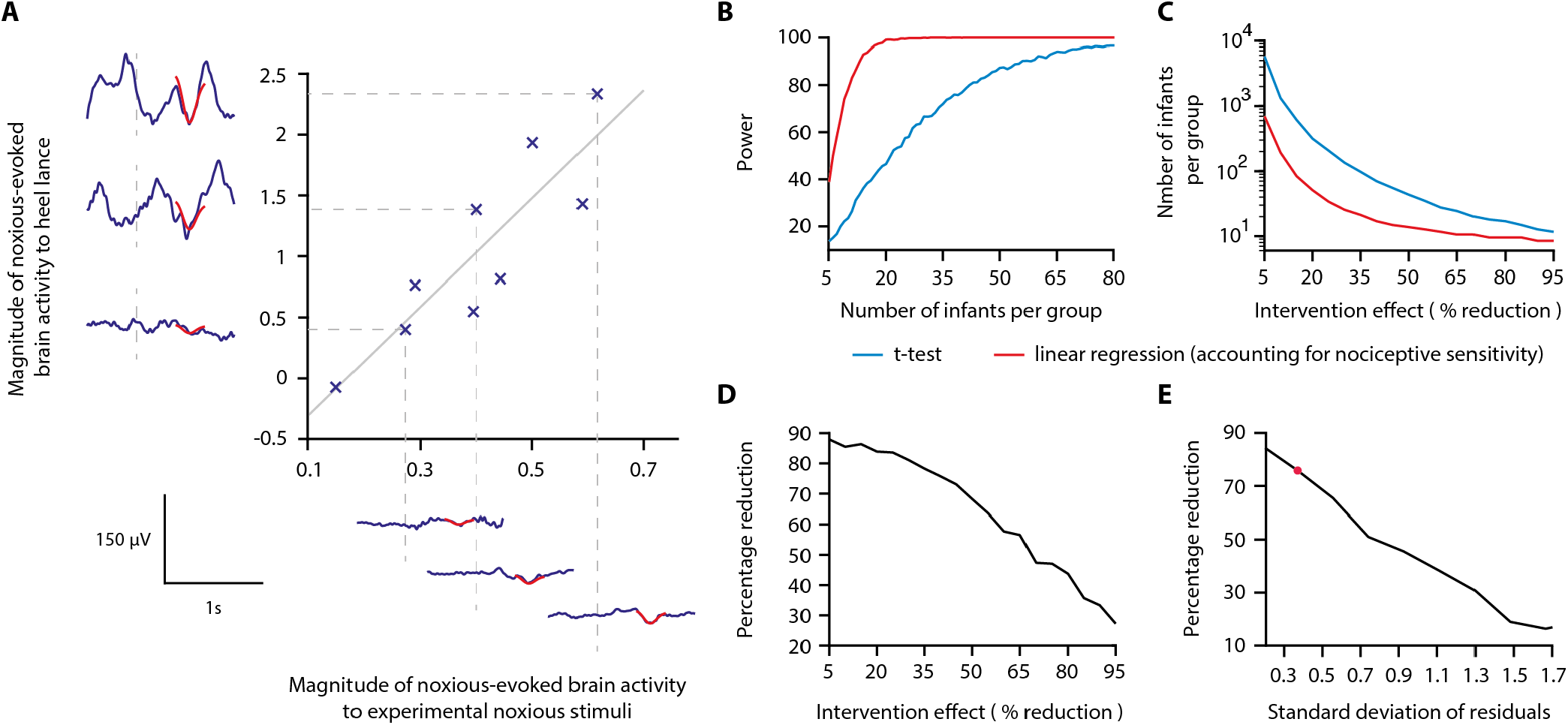
Magnitude of noxious-evoked brain activity in response to experimental noxious stimuli correlates with the response to a clinically-required heel lance and can be used as a measure of nociceptive sensitivity to reduce sample sizes. **(A)** The magnitude of noxious-evoked brain activity following mild experimental noxious stimuli and a clinically-required heel lance was significantly correlated within-subject (p = 0.0025, R^2^ = 0.77, n=9, Pearson correlation test, Study 1); grey solid line indicates line of best fit. Dashed lines and their corresponding EEG traces indicate three infants with a range of response magnitudes. The magnitude of the brain activity was quantified using a template of noxious-evoked activity, shown overlaid in red (Hartley et al., 2017). **(B-E)** In Study 2 we used simulated data to investigate how sample size is altered when the relationship in (A) is considered. **(B)** For each sample size 1000 data sets were simulated with a 40% reduction in the response to a clinically-required procedure assumed in the Intervention Group. The power (percentage of significant results, p<0.05) to detect a difference between the two groups was calculated for each sample size using a linear regression with (red) and without (blue) accounting for individual differences in nociceptive sensitivity. **(C)** The number of infants required to achieve 95% power with different levels of intervention effect. Simulations were run with increasing numbers of infants until 95% power was achieved. **(D)** Percentage reduction in the number of infants required per group when individual nociceptive sensitivity is accounted for compared with not accounting for nociceptive sensitivity (power=95%). **(E)** The percentage reduction in the number of infants required per group with different degrees of correlation (standard deviation of residuals) between the responses to experimental indicates the standard deviation of residuals (SD=0.37) in (A).

### Study 2: Simulating the effect of accounting for individual nociceptive sensitivity

In Study 2, we used simulated data to investigate whether accounting for individual differences in baseline nociceptive sensitivity has the potential to reduce the sample size needed to assess the efficacy of an analgesic intervention. Here we initially assume that an effective analgesic intervention results in a 40% reduction in noxious-evoked brain activity; this is clinically meaningful as a similar reduction in noxious-evoked brain activity is observed when adults report significantly lower verbal pain scores (Lorenz et al., 1997; von Mohr et al., 2018). We simulated an Intervention Group and Control Group across a range of sample sizes, simulating both baseline nociceptive sensitivity data and responses to heel lance, and assuming the relationship between these measures observed in Study 1 is maintained.

The simulated Control Group and the Intervention Group responses were compared using a linear regression with and without baseline nociceptive sensitivity as a covariate (see Methods). At a significance level of 0.05, the sample size to achieve a given power is substantially reduced when nociceptive sensitivity is accounted for (Figure 2B). The reduction in sample size that can be achieved by accounting for individual differences in nociceptive sensitivity is highly dependent on the anticipated effect size of the intervention (Figure 2C, D). For example, at the extremes we considered, with an assumed intervention effect size of 95% (and power of 95%) a sample of 11 infants per group would be required without accounting for nociceptive sensitivity compared with eight infants per group when nociceptive sensitivity is accounted for, a 27% reduction in sample size. Whereas, by comparison, assuming an intervention effect size of 5%, the sample size required to achieve 95% power is 5458 infants per group without accounting for individual nociceptive sensitivity, compared with 660 infants per group when nociceptive sensitivity is accounted for – representing an 88% reduction in sample size (Figure 2C, D). Assuming an intervention effect size of 40%, a sample size of 16 infants per group (32 infants in total) would be sufficient to observe a significant intervention effect with 95% power if individual differences in nociceptive sensitivity are accounted for. In contrast, a sample size of 66 infants per group (132 infants in total) is required to achieve the same power if infant nociceptive sensitivity is not accounted for (Figure 2B).

The percentage reduction in sample size which can be achieved by accounting for individual differences in nociceptive sensitivity is also dependent on the strength of the correlation between the brain responses evoked by the experimental stimuli and the acute clinical procedure (Figure 2E). When a weak correlation exists between the measures (calculated using the standard deviation of the correlation residuals), the reduction in sample size is low. Conversely, with a strong correlation between measures, a greater reduction in sample size is achieved. For example, with an assumed intervention effect of 40% and the low standard deviation of the residuals observed in study 1 (SD of residuals =0.37), accounting for nociceptive sensitivity results in a sample size reduction of approximately 76%, compared to a sample size reduction of 17% when a high noise level (SD of residuals =1.7) is observed in the correlation (Figure 2E).

### Study 3: Testing the paradigm: a non-pharmacological pain-relieving intervention study

In a previous study, we reported that a non-pharmacological gentle touch intervention, (brushing an infant’s leg at a rate of approximately 3cm/s to optimally stimulate C-tactile fibres) prior to a clinically-required heel lance caused a 40% reduction in noxious-evoked brain activity (Gursul et al., 2018). In Study 3, we used the same non-pharmacological intervention in an independent prospective cohort of healthy infants that clinically required a heel lance for the purpose of blood sampling and tested the effect of incorporating the nociceptive sensitivity paradigm and accounting for inter-individual differences in baseline nociceptive sensitivity. Based on power calculations from simulated data in Study 2, assuming a 40% reduction in noxious-evoked brain activity from the intervention and 95% power, a total of 16 infants were included in the Intervention Group and were gently brushed on the leg ipsilateral to the stimulus site at a rate of approximately 3cm/s for 10 seconds prior to heel lancing (Gursul et al., 2018). A further 15 infants were included in the Control Group where the heel lance was performed without gentle brushing. All infants received mild experimental noxious stimulation prior to heel lancing to assess their individual baseline nociceptive sensitivity (see Methods). Unlike Study 1, in which infants had been stimulated with a force of 64 mN, a force of 128mN was applied in this prospective cohort to increase the signal-to-noise ratio. The necessary strong correlation between the evoked response to the experimental stimulus and clinical procedure was confirmed in the Control group (p=0.0013, R^2^=0.65, Figure 3A).

**Fig. 3.**
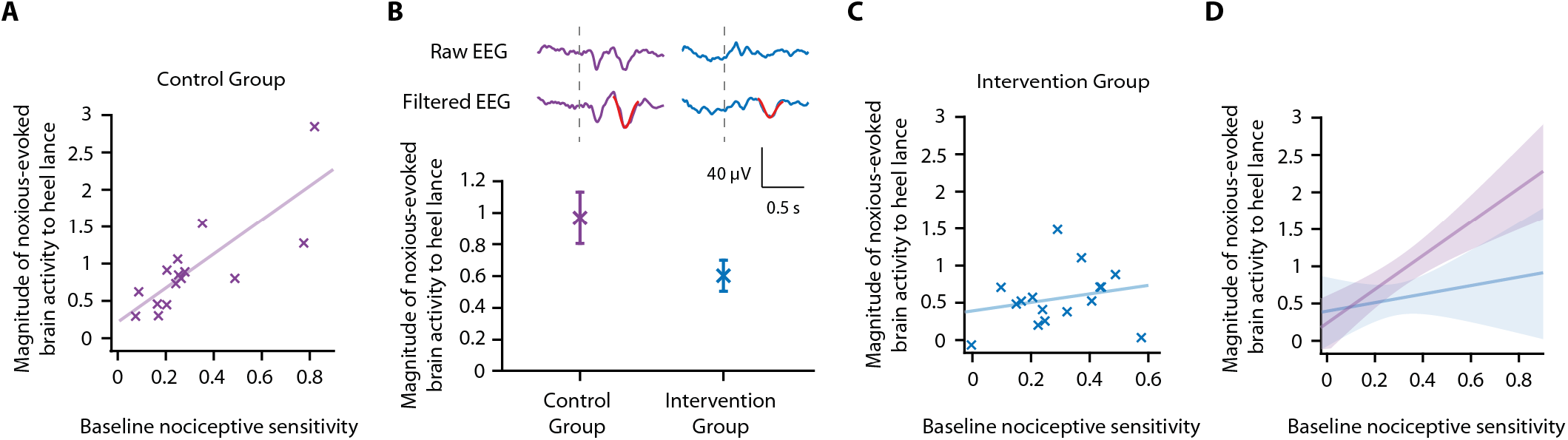
Accounting for individual nociceptive sensitivity in the assessment of efficacy of a gentle touch intervention. **(A)** The magnitude of the noxious-evoked brain activity following a mild experimental noxious stimulus compared with the clinically-required heel lance for each infant in the Control Group (n=15). Solid line indicates line of best fit. **(B)** (Top) Group average raw EEG and (Woody) filtered EEG traces in response to the clinically-required heel lance; Control Group (purple) and Intervention Group (infants received gentle touch at a rate of approximately 3cm/s for 10 seconds prior to the heel lance, n=16) (blue). Dashed lines indicate the point of stimulation; the template of noxious-evoked brain activity is shown overlaid in red. (Bottom) Magnitude of the noxious-evoked brain activity following heel lance in the two groups. Error bars indicate mean ± standard error. **(C)** Comparison of the stimulus responses for each infant in the Intervention Group. Gentle touch was not applied prior to the experimental noxious stimuli so that each infant’s individual baseline nociceptive sensitivity could be assessed. **(D)** Confidence intervals of the correlations for the two groups shown overlaid: Control Group (purple), Intervention Group (blue), solid lines indicate line of best fit. The effect of the intervention (gentle touch) is demonstrated by the difference between the two groups’ confidence intervals and is most evident in infants who have greater baseline nociceptive sensitivity.

Consistent with the previously published study (Gursul et al., 2018), the gentle touch intervention resulted in a 39% reduction in the magnitude of the noxious-evoked brain activity, but a significant intervention effect was not observed (although the result indicated borderline significance) when nociceptive sensitivity was not accounted for, likely due to the lack of power with this sample size (linear regression, t=1.95, p=0.05, Figure 3B, Study 2 indicates a power of 40% for a sample of this size without accounting for nociceptive sensitivity, Figure 2B). However, when baseline nociceptive sensitivity was accounted for as a covariate in the analysis, a significant intervention effect was observed (linear regression, t=2.29, p=0.026).

To further understand these results we compared the relationship between the responses within the Control Group and Intervention Group. Unlike the significant correlation between the magnitude of noxious-evoked brain activity in response to the experimental noxious stimuli and heel lancing demonstrated in the Control group (p=0.0013, R^2^=0.65, Figure 3A), this relationship was disrupted in the Intervention Group (p=0.39, R^2^=0.05, Figure 3C). In particular, we observed reduced noxious-evoked brain activity following the gentle brushing intervention in infants with high baseline nociceptive sensitivity (Figure 3D), suggesting that the effect of pain-relieving interventions is most prominent in infants with greater baseline nociceptive sensitivity. Each infant’s EEG responses to the experimental noxious stimulus and the heel lance are shown in Figure S1.

### Testing the paradigm with other modalities: noxious-evoked reflex withdrawal

Noxious stimulation in infants evokes a range of physiological responses including facial grimacing, reflex withdrawal and physiological responses (Cornelissen et al., 2013; Hartley et al., 2015; Hatfield and Ely, 2015). There is great value in establishing whether accounting for individual differences in nociceptive sensitivity can be applied to other pain-related measures. In Study 3, the magnitude of the reflex withdrawal was also recorded in response to the experimental noxious stimulation and heel lancing. In the Control Group, the magnitude of the reflex withdrawal response to experimental noxious stimulation was significantly correlated with the reflex withdrawal evoked by heel lancing (p=0.009, R^2^=0.36, Figure 4A). However, this correlation in reflex withdrawal activity was weaker than the relationship in the noxious-evoked brain activity, limiting its use (Figure 2E). Assuming an intervention effect of 40% and this level of correlation identified within the same size sample, simulated data reveals that accounting for nociceptive sensitivity using reflex activity provides only 17.3% power to detect a significant difference between the two groups compared with a power of 11.3% without accounting for nociceptive sensitivity.

**Fig. 4.**
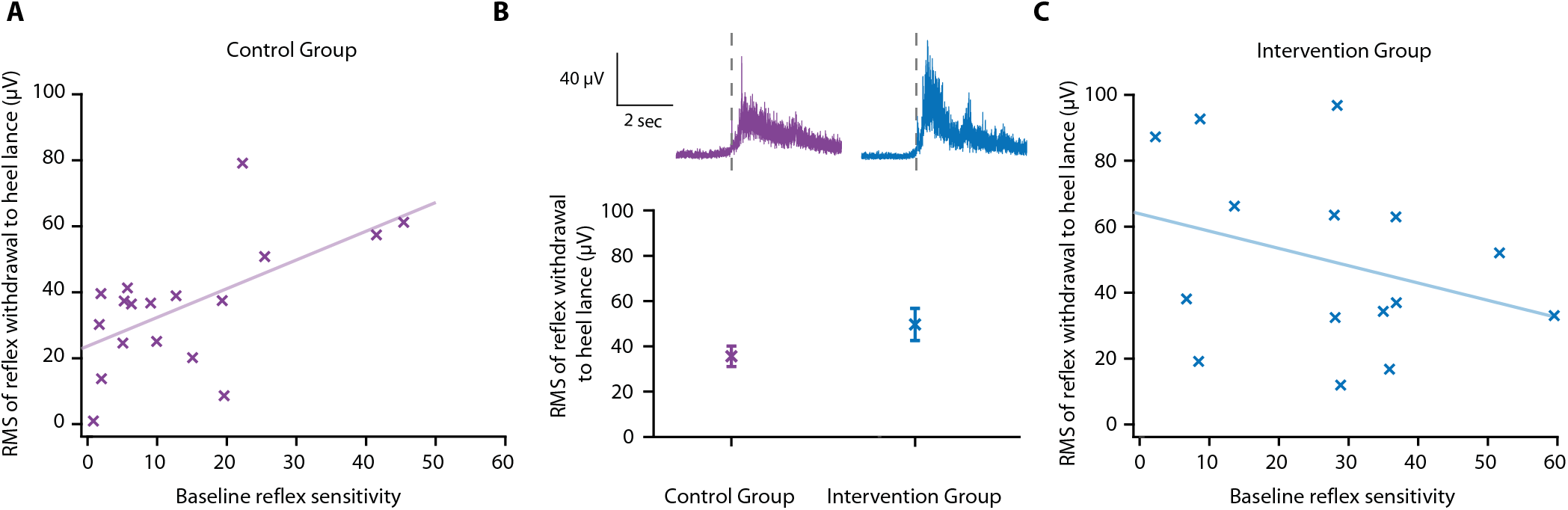
Effect of gentle touch on reflex responses. **(A)** The magnitude of the reflex withdrawal following a mild experimental noxious stimulus (baseline reflex sensitivity) compared with the clinically-required heel lance for each infant in the Control Group (n=18). Solid line indicates line of best fit. **(B)** Average EMG traces (top) for infants in the Control Group (purple) and Intervention Group (blue, n=15) where infants were gently brushed at a rate of approximately 3cm/s for 10 seconds prior to the heel lance. Dashed lines indicate the point of stimulation. (Bottom) Magnitude of the reflex withdrawal response in the two groups. Error bars indicate mean of the root mean square (RMS) of the reflex withdrawal ± standard error. **(C)** The magnitude of the reflex withdrawal following a mild experimental noxious stimulus (baseline reflex sensitivity) compared with the clinically-required heel lance for each infant in the Intervention Group (gentle touch).

In this study, the gentle touch intervention did not significantly reduce the magnitude of the reflex withdrawal activity following heel lancing, either when accounting for nociceptive sensitivity (linear regression, t=-1.43, p=0.17,) or without accounting for nociceptive sensitivity (t=-1.73, p=0.10, Figure 4B). While it is possible that reflex withdrawal of the stimulated limb is not modulated by gentle touch, as has previously been suggested (Gursul et al., 2018), the intervention clearly disrupted the correlation between baseline reflex sensitivity and the reflex evoked by heel lancing (p=0.25, R^2^=0.1, Figure 4C). The brushing intervention may have caused a change in baseline muscle activity in some individuals resulting in the larger residuals in the Intervention Group (SD of the residuals – Control Group: 15.3 µV, Intervention Group: 26.2 µV). Each infant’s EMG responses to the experimental noxious stimulus and the heel lance are shown in Figure S2.

### Study 4: A pharmacological analgesic study

In Study 4, we conducted an opportunistic study to investigate whether the administration of paracetamol prior to immunisation significantly reduces noxious-evoked brain activity. In 2015, national clinical guidelines recommended the administration of paracetamol at the time of Meningitis B immunisation due to its antipyretic effect (Public Health England and NHS England, 2015). Therefore, our local neonatal unit (John Radcliffe Hospital) began administering oral paracetamol to infants immediately after vaccination. In October 2018, the local practice guidelines were updated, recommending the administration of oral paracetamol one-hour pre-vaccination. Prior to the guideline change, we studied 17 infants who did not receive paracetamol before immunisations (Control Group), recording their noxious-evoked brain activity during immunisations. Following the guideline change, we recorded noxious-evoked brain activity in 16 infants who received paracetamol one-hour prior to immunisations (Intervention Group) (see Methods and Figure 5A). In the Intervention Group we explored the relationship between baseline nociceptive sensitivity and brain activity evoked by immunisation following paracetamol administration.

**Fig. 5.**
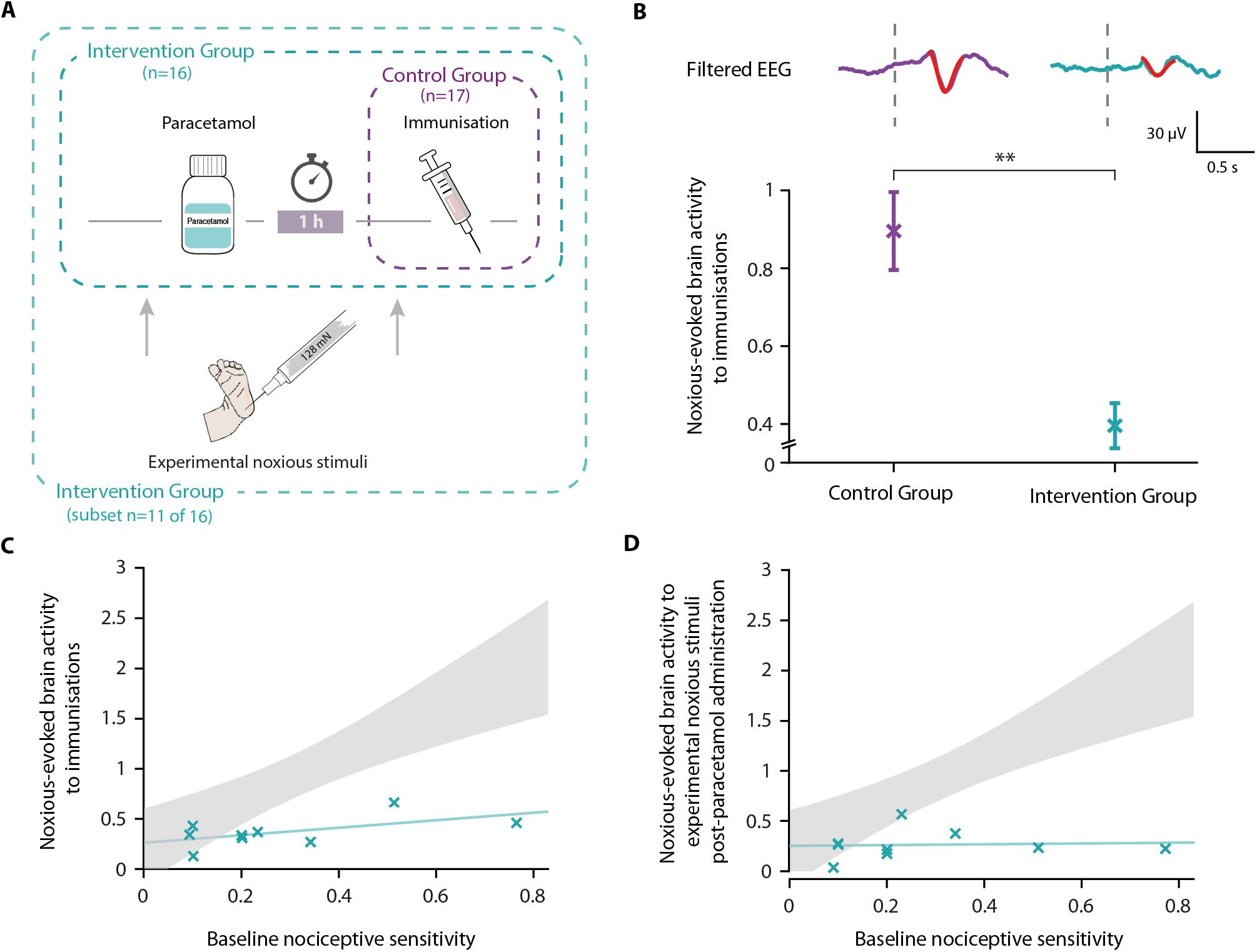
Paracetamol significantly reduces noxious-evoked brain activity following immunisation. **(A)** Experimental design of Study 4. EEG was recorded during immunisations in infants before (Control Group, n=17) and after the guideline change (Intervention Group, n=16, received paracetamol one-hour prior to immunisation). A subset of infants in the Intervention Group (n=11 of 16) received experimental noxious stimuli before and approximately one-hour after paracetamol administration. **(B)** Average (Woody) filtered EEG following immunisations are displayed (top); for the Control Group in purple and Intervention Group in teal, the template of noxious-evoked brain activity is shown overlaid in red. Dashed lines indicate the point of contact of the needle with the skin. (Bottom) Magnitude of the noxious-evoked brain activity following immunisations in the two groups, error bars indicate mean ± standard error (linear mixed effects regression model - without accounting for nociceptive sensitivity, **p<0.001). **(C)** Magnitude of the noxious-evoked brain activity following the experimental noxious stimulus prior to paracetamol administration (baseline nociceptive sensitivity) compared with the noxious-evoked brain activity to immunisation (which was approximately one-hour after paracetamol administration) for each infant in the Intervention Group subset (markers in teal). For comparison, the confidence interval of the Control Group correlation in Study 3 (i.e. the correlation between the response to experimental noxious stimuli and a heel lance) is shown in grey. **(D)** Magnitude of the noxious-evoked brain activity to experimental noxious stimuli prior to paracetamol administration (baseline nociceptive sensitivity) compared with the noxious-evoked brain activity to experimental noxious stimuli applied approximately one-hour after paracetamol administration for each infant in the Intervention Group subset (markers in teal). For comparison, the confidence interval of the Control Group correlation in Study 3 (i.e. the correlation between the response to experimental noxious stimuli and a heel lance) is shown in grey.

Noxious-evoked brain activity in response to immunisation was characterised using a fast frame rate video camera to identify the time when the needle first came into contact with the skin (Hartley et al., 2014; Verriotis et al., 2015). For each infant, up to 3 immunisations were recorded on the same test occasion. First, we validated that the template of noxious-evoked brain activity (Hartley et al., 2017) can be used to quantify the magnitude of noxious-evoked brain activity from immunisation applied to the thigh (experimental data presented in Supplementary Materials, Figure S3 and S4). The magnitude of noxious-evoked brain activity following immunisation was significantly lower in the infants who received paracetamol prior to vaccination (linear mixed effects regression model with subject and number of immunisations set as random effects, Control Group mean: 0.88 [SD 0.58] (n=15); Intervention Group mean: 0.40 [SD 0.30] (n=14), t=3.61, p<0.001, Figure 4B). Individual infant’s EEG responses to the immunisation are shown in Figure S5.

In a subset of 11 of the 16 infants in the Intervention Group, who received paracetamol prior to immunisation, we also recorded responses to experimental noxious stimulation before and approximately one-hour after paracetamol administration (Intervention Group subset, Figure 5A). As this study was implemented opportunistically following changes in clinical guidelines, responses to experimental noxious stimuli were not recorded in all infants. Nevertheless, the baseline nociceptive sensitivity measures that were recorded in response to the experimental noxious stimuli applied prior to paracetamol administration, had a range of values that were similar to Studies 1 and 3 (range: 0.09 – 0.77). Likewise, the magnitude of the brain activity evoked by the immunisations were similar to that evoked by heel lance in the previous studies for both the Intervention Group (Intervention Group (Study 3) range: −0.06 – 1.48; Intervention Group (Study 4) range: −0.08 – 1.22) and the Control Group (Control Group (Study 3) range: 0.30 – 2.84; Control Group (Study 4) range: −0.07 – 2.14). Although we did not record the baseline sensitivity in infants in the Control Group, in the absence of a pain-relieving intervention we would expect the response to be correlated with noxious-evoked brain activity evoked by immunisation. As the correlation between baseline sensitivity and response to immunisation was low in the Intervention Group (p=0.12, R^2^=0.33, n=9, Figure 5C), the relationship between these measures was likely disrupted by paracetamol. Similar to the gentle brush intervention, the infants with high baseline nociceptive sensitivity, represented by a high magnitude response to experimental noxious stimulation prior to paracetamol administration, had much lower magnitude responses to immunisation than would have been expected without an analgesic intervention (Figure 5C). Similarly, the correlation between the baseline nociceptive sensitivity (magnitude of the baseline nociceptive sensitivity prior to paracetamol administration) and the response to the experimental noxious stimuli one-hour post-paracetamol administration was disrupted (p=0.83, R^2^=0.006, n=9, Figure 5D). There was no significant difference in the responses to the experimental noxious stimuli before and after paracetamol administration (linear regression, before paracetamol mean: 0.27 [SD 0.38]; after paracetamol mean: 0.27 [SD 0.35], t=0.17, p=0.86, n=9) but we were likely not powered to observe an effect.

## Discussion

We demonstrate that accounting for individual differences in nociceptive sensitivity significantly reduces the sample size required to assess the efficacy of analgesics in infants. Noxious-evoked brain activity in response to a low-intensity experimental noxious stimulus can be used in infants as a marker of baseline nociceptive sensitivity and is highly correlated with the magnitude of noxious-evoked brain activity produced by clinically-required acute painful procedures. Using both simulated and experimental data, we demonstrate that the sample size required to observe the effects of analgesic interventions (for a given power and significance level) can be significantly reduced when nociceptive sensitivity is accounted for. Importantly, the percentage reduction in sample size is related to the expected effect size of the intervention and the degree of correlation between the baseline nociceptive sensitivity measure and the brain activity evoked by the clinical procedure. By testing this novel paradigm in clinical studies, we re-confirm the efficacy of gentle touch as a non-pharmacological intervention that reduces brain activity evoked by heel lancing (Gursul et al., 2018) and we provide evidence to suggest that oral paracetamol is a candidate analgesic drug for procedural pain in infants. Although these studies have a number of limitations (including lack of randomisation), and only investigate one aspect of the infant response to nociceptive input (namely an EEG-derived noxious-evoked potential), they provide strong evidence to suggest that randomised clinical trials investigating the efficacy of both gentle touch and paracetamol through multimodal pain assessment measures are warranted.

Minimisation of sample sizes is imperative in clinical research, and particularly in neonatal studies given the inherent ethical, recruitment, and experimental challenges associated with studying this patient population. Considering that inter-individual variability drives increases in sample sizes required to demonstrate efficacy, addressing baseline variability is key. The novel experimental paradigm we present provides a robust approach to indirectly control for a vast array of known and unknown demographic and environmental factors that influence nociception and result in inter-individual variability in responses, as well as potential experimental confounds which differ between individuals (such as differences in signal-to-noise ratios, head circumference and skull thickness). The experimental noxious stimulus used in these studies provides a practical and ethical paradigm for the assessment of baseline nociceptive sensitivity in infants. It is non-tissue damaging in both term and ex-premature infants, activates A*∂* and C fibres (van den Broeke et al., 2015), does not evoke changes in facial expression or signs of behavioural distress (Goksan et al., 2015; Hartley et al., 2017, 2015), and is acceptable to parents. The application of experimental noxious stimuli provides a reliable measure of baseline nociceptive sensitivity as the mild stimuli can be repeated and trial averages calculated within individual infants; an approach that substantially reduces the signal-to-noise ratio as compared with responses recorded in response to a single clinical procedure. Moreover, there is no evidence from our data that the experimental noxious stimuli increases the magnitude of the heel lance response given that the responses to heel lance reported here are similar to previous papers where the experimental noxious stimuli was not applied (Hartley et al., 2017), suggesting it is appropriate for use in a clinical setting. Despite the advantage of using this approach, we cannot rule out the potential effects of selection bias (Bishop, 2020). A relatively high number of trials were rejected due to artefacts, which may be more pronounced when there are stimulus-related movements. If these movements are indicative of a more vigorous response to the nociceptive input, then it is plausible that we are unavoidably biased towards a subset of the population.

The applicability of the nociceptive sensitivity paradigm was tested in the context of a pain-relieving intervention that we have previously shown to be effective in reducing noxious-evoked brain activity – gentle touch (Gursul et al., 2018). Infants were gently brushed at a speed of 3 cm/s, which is approximately equivalent to the rate at which parents will naturally stroke their infants (Croy et al., 2016) and optimises stimulation of C-tactile fibres (Löken et al., 2009). In an independent population of infants, we re-confirmed that brushing the skin prior to a clinically-required heel lance significantly reduces noxious-evoked brain activity. We used our nociceptive sensitivity paradigm to indirectly account for many factors that influence the magnitude of noxious-evoked brain activity. In addition, we did not observe a significant difference in reflex withdrawal activity between the control infants and the infants who received gentle touch prior to the heel lance, which is consistent with our previous observations (Gursul et al., 2018). It is possible that either the magnitude of the reflex withdrawal is genuinely not modulated by the brush intervention or that a modulation in reflex activity would only be observed with a larger sample size. Importantly, a significant but weak correlation was observed between the reflex activity in response to the noxious stimuli and in response to heel lancing in the Control Group, suggesting that the paradigm presented here could be useful in future trials where reflex withdrawal activity is used as an outcome measure.

In addition to minimising sample sizes, assessing baseline nociceptive sensitivity may also allow for identification of infants that would benefit most from analgesic interventions. Infants with larger baseline nociceptive sensitivity had the greatest reduction in response following the intervention. In contrast, infants with low baseline nociceptive sensitivity were less likely to demonstrate a benefit of the intervention, as for this clinical procedure the potential reduction in their responses was minimal. This could be due to a floor effect whereby for some infants noxious-evoked brain responses to heel lance is close to zero and cannot be reduced further. Improving our understanding of inter-individual variability in pain responses is pivotal to ensure that for each individual infant potential adverse effects of analgesics are carefully weighed against potential benefits.

In our final study, we demonstrate that paracetamol significantly reduced the magnitude of the noxious-evoked brain activity following immunisation compared with infants who did not receive paracetamol prior to immunisation. Although this result is consistent with studies in adults, using both EEG (Bromm et al., 1992; Pickering et al., 2013) and fMRI (Pickering et al., 2015), where paracetamol has been shown to reduce brain activity evoked by nociceptive procedures, previous studies in infants have provided insufficient evidence to determine the analgesic efficacy of paracetamol for acute procedural pain (Ohlsson and Shah, 2020). While several studies suggest an opioid-sparing effect of paracetamol (Ceelie et al., 2013; Härmä et al., 2016), the majority of studies do not demonstrate a reduction in behavioural or physiological responses to commonly performed painful procedures, such as heel lancing (Badiee and Torcan, 2009; Bonetto et al., 2008; Shah et al., 1998) and invasive eye examinations to screen for retinopathy of prematurity (Seifi et al., 2013). The behavioural outcome measures used in these studies may fail to discriminate between pain and distress (Moultrie et al., 2017; Slater, 2019), which could limit conclusions related to analgesic efficacy. However, given the small sample size of the present study and that we are only characterising the immediate nociceptive brain activity evoked by the needle insertion, rather than the pain associated with the injection of the fluid into the muscle for example; randomised clinical trials that also include other pain-related measures are warranted to assess the benefit of paracetamol administration prior to immunisation. Nonetheless, small studies in adults demonstrate that candidate drugs can modulate pain-related neural activity in the absence of verbally-reported analgesia, and these brain-derived measures are recognised as a valuable approach to obtain objective evidence related to potential analgesic efficacy in early proof of concept studies (Wanigasekera et al., 2018). The noxious-evoked brain activity measure used here quantifies the evoked potential produced at the central vertex electrode site (Cz), which has been shown to have the greatest and most reproducible response size amplitude compared to other electrodes sites across the brain (Hartley et al., 2017; Verriotis et al., 2015). This measure does not represent all nociceptive activity across the brain and cannot be used to investigate the various aspects of pain perception (Hartley et al., 2017); a multi-modal approach to pain assessment is therefore important in follow-on studies (Vaart et al., 2019). However, in the absence of verbalisation, neuroimaging methods provide an objective proxy approach which has been used to infer pain perception following noxious events (Baxter et al., n.d.; Duff et al., 2020; Gursul et al., 2019; Hartley et al., 2017).

Paracetamol is administered as an antipyretic for the Meningitis B immunisations. An update to our local clinical guidelines was implemented, whereby the paracetamol was administered prior to rather than after immunisation. This meant we were able to opportunistically study whether paracetamol can reduce noxious-evoked brain activity following immunisation. Nevertheless, our study is significantly limited due to the restricted sample sizes, lack of randomisation and blinding, and because in the Control Group, where paracetamol was administered after immunisation, we did not record baseline nociceptive sensitivity prior to immunisation. Therefore, we do not have data to confirm that the baseline nociceptive sensitivity is correlated with the magnitude of the evoked brain activity following immunisation; although, given there is no discernible correlation between these measures in the Intervention Group, this strongly suggests that this relationship has been disrupted by paracetamol administration. Furthermore, for infants with high baseline nociceptive sensitivity, the brain responses evoked by the immunisation were much lower than would be expected in the absence of the analgesic intervention. To broaden the utility of this paradigm, it will be necessary to characterise the correlation between baseline nociceptive sensitivity and a range of acute clinical procedures, including immunisation.

Although many factors that influence individual variability in pain sensitivity in infants are accounted for using the nociceptive sensitivity paradigm, it does not account for differences in rapidly fluctuating trait effects such as differences in attention or sleep state that could vary between the baseline sensitivity testing and the implementation of the clinical procedure. Understanding how trait differences influence variability in pain responses will facilitate better estimation of the expected responses to clinically-required painful procedures. A recent fMRI study demonstrated that noxious-evoked brain activity can be predicted from an infant’s resting state brain activity as well as the structural integrity of key white matter pathways (Baxter et al., n.d.). Investigating the role of baseline EEG activity and exploring the neurological differences underlying variability in the pain-related brain activity described here could further improve the utility of the paradigm.

In summary, the assessment of pain in non-verbal infants is challenging (Slater, 2019) and the wide variability in individual responses to painful procedures complicates the assessment of analgesics. Currently there is a paucity of evidence regarding the efficacy of pain-relieving interventions used in neonatal practice (Neville et al., 2014). Here, we present a paradigm that accounts for individual nociceptive sensitivity and we demonstrate its utility in terms of sample size reduction. Using this paradigm in clinical trials could optimise resources, maximise the value of collected data and ultimately expedite the discovery and validation of urgently needed analgesics for this patient population.

## Methods

### Study design and participants

A total of 99 infants were included in four studies. In Study 1, the relationship between responses to experimental noxious stimuli and clinically-required heel lance was investigated in nine infants using data previously collected for other (unpublished) studies. In Study 2, the potential value of the statistical relationship identified in Study 1 was investigated using computational simulations. In Study 3, brain activity and reflex withdrawal responses from 40 infants were recorded to test the paradigm with gentle touch as a pain-relief intervention. In Study 4 brain activity was recorded from 33 infants in response to immunisations to test the analgesic efficacy of paracetamol. Additionally, the brain-derived measures to characterise immunisation evoked activity were validated in a further 17 infants (Supplementary Material).

The participants were recruited from the Maternity Unit and Newborn Care Unit at the John Radcliffe Hospital, Oxford University Hospitals National Health Service Foundation Trust, Oxford, UK. Medical charts were reviewed, and infants assessed as clinically stable, not receiving analgesics at the time of study (except from paracetamol where specified) and with no history of neurological problems or maternal substance abuse were eligible for inclusion. Infant demographics are presented in Table 1.

**Table 1.** Infant demographics. Values given are median (lower quartile, upper quartile) or number (%). * Indicates a missing score for one infant transferred to NICU from another hospital after birth.

|  | Study 1 | Study 3 |  | Study 4 |  |  |
| --- | --- | --- | --- | --- | --- | --- |
|  |  | Control Group | Intervention Group | Control Group | Intervention Group | Template validation (Supplementary Materials) |
| Number of infants | 9 | 18 | 22 | 17 | 16 | 17 |
| Gestational age (GA) at birth (weeks) | 40.7 (40.3, 41) | 40 (37.1, 40.7) | 39.2 (37.2, 40.9) | 28.1 (26, 28.9) | 28 (26.4, 28.3) | 40.6 (39.9, 41) |
| Postmenstrual age (PMA) at time of study (weeks) | 41 (40.9, 41.7) | 40.5 (37.6, 40.9) | 39.9 (37.9, 41.4) | 38 (37.1, 38.6) | 37 (36.1, 38.1) | 40.7 (40.1, 41.4) |
| Postnatal age (PNA) at time of study (days) | 2 (2, 4) | 1.5 (1, 3.5) | 3.5 (2, 5) | 64 (58, 82) | 64 (60.5, 70.3) | 2 (1, 3) |
| Birthweight (g) | 4140 (3725, 4320) | 3675 (3021, 3881) | 3535 (3101, 4080) | 1120 (790, 1365) | 910 (743, 1054) | 3480 (3130, 3730) |
| Sex |  |  |  |  |  |  |
| Male | 4 (44) | 11 (61) | 10 (45.5) | 11 (65) | 10 (62.5) | 13 (76) |
| Female | 5 (56) | 7 (39) | 12 (54.5) | 6 (35) | 6 (37.5) | 4 (24) |
| Mode of delivery |  |  |  |  |  |  |
| NVD | 2 (22) | 6 (33) | 8 (36) | 7 (41) | 3 (19) | 8 (47) |
| Assisted vaginal ventouse/forceps | 6 (67) | 3 (17) | 4 (18) | 0 (0) | 1 (6) | 5 (29) |
| Emergency C-Section | 0 (0) | 6 (33) | 5 (23) | 8 (47) | 9 (56) | 4 (24) |
| Elective C-Section | 1 (11) | 3 (17) | 5 (23) | 2 (12) | 3 (19) | 0 (0) |
| Apgar score at 1 min | 9 (8, 10) | 9 (8, 10) | 9 (7, 10) | 5 (4, 6) * | 7 (3, 8) * | 9 (9, 10) |
| Apgar score at 5 min | 10 (10, 10) | 10 (10, 10) | 10 (9, 10) | 8 (7, 10) * | 8 (7, 10) * | 10 (10, 10) |

### Research governance

Studies were conducted in accordance with the Declaration of Helsinki and Good Clinical Practice guidelines. Ethical approval was obtained from the National Research Ethics Service (reference 12/SC/0447) and informed written parental consent was obtained prior to each study.

## Experimental design

### Study 1. Characterising individual baseline nociceptive sensitivity using brain activity in infants

The aim of this study was to investigate the relationship between noxious-evoked brain activity in response to experimental stimuli and clinically-required heel lance within-subjects in a group of term infants. We retrospectively searched all the data we had previously collected (and that has not been previously published) to identify any term infants who had received a clinically-required heel lance and experimental noxious stimuli on the same test occasion. We identified 9 infants studied between 2014 and 2015 (age range: 39 to 42 weeks’ gestational age (GA)) who had all received experimental noxious stimuli at a force of 64 mN. The magnitude of the noxious-evoked brain activity was characterised by projecting a previously described template of noxious-evoked brain activity (Hartley et al., 2017) (see *Recording techniques and analysis* for further details) in response to each stimulus. The mean response to the experimental noxious stimulus in each infant was correlated with their responses to the heel lance using a Pearson correlation test.

### Study 2: Simulating the effect of accounting for individual nociceptive sensitivity to reduce sample sizes

To investigate potential differences in the power achieved by accounting for individual nociceptive sensitivity at different sample sizes (for a given effect size and significance level) we simulated data sets. Each simulation consisted of a control group and an intervention group. Given a sample size of N per group, we first simulated N individual nociceptive sensitivity levels for each group by generating N uniform random numbers within the range of expected sensitivities. The minimum expected sensitivity was set as the minimum response to the experimental noxious stimuli in the data collected in Study 1, and the maximum expected response was set from multiplying the maximum of data collected in Study 1 by 3 (the expected increase in range from changing to an experimental noxious stimulus with a force of 128 mN from previously published data - Study 2 in (Hartley et al., 2017), as using a force of 128 mN is expected to increase the signal-to-noise ratio). Thus, N individual nociceptive sensitivities, *x*_*i*_, were generated per group with

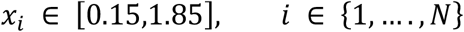

Responses to the clinical stimulus, *y*_*i*_, were then simulated from these randomly generated individual nociceptive sensitives with:

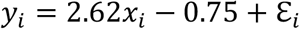

Where ε_*i*_ is a noise term, and the values 2.62 and 0.75 were related to the line of best fit in Study 1 (the gradient of the line was reduced from Study 1 to account for the increase in the range of individual nociceptive sensitivities, i.e. as the range of *x*_*i*_ was increased from Study 1 the gradient was reduced so that the maximum value of *y*_*i*_ was not higher than the expected response magnitude to a heel lance).

ε_*i*_ was drawn from a random normal distribution with mean of 0 and a standard deviation of ξ. The standard deviation of residuals ξ was set to be 0.37 in most simulations as this is the standard deviation of the residuals from Study 1 but was varied for the simulations in Figure 2E.

Finally, the responses to the clinical stimulus in the intervention group were reduced by a proportion according to the intervention effect. For most simulations the intervention effect was set at 40% as this is considered a clinically meaningful effect size (Lorenz et al., 1997; von Mohr et al., 2018). However, varying levels of intervention effects were also investigated. We compared the responses in the control group and the intervention group with and without accounting for nociceptive sensitivity (see Statistical Analysis).

For each value of N, 1000 control and intervention groups were simulated and the percentage of simulations where the group comparisons had p < 0.05, i.e. the power, was calculated. For simulations where the intervention effect or the noise level were varied, the minimum number of infants required for a power of 95% was calculated by increasing the group size by 1 (note all data was re-simulated with each new sample size – so each simulated data set was fully independent) and calculating the power at each step until a power of 95% was achieved.

### Study 3: Testing the paradigm: a non-pharmacological pain-relieving intervention study

The aim of this study was to test the nociceptive sensitivity paradigm using a gentle touch intervention of known effect in reducing the noxious-evoked brain activity following a clinically-required heel lance (Gursul et al., 2018). The sample size required was obtained from the data simulated in Study 2. Assuming a 40% reduction in the magnitude of the brain activity in the Intervention Group compared with the Control Group, a sample size of 32 infants (16/group) would be sufficient to achieve a power of 95%, when an infant’s baseline nociceptive sensitivity is taken into account. A total of 40 infants aged 35 to 42 weeks PMA were prospectively recruited to the study. EEG and EMG activity were recorded in response to experimental noxious stimuli (128 mN intensity) prior to a clinically-required heel lance. The Intervention Group (n=22) received gentle touch at an approximate rate of 3cm/s for 10 seconds before heel lancing and the Control Group (n=18) did not receive the gentle touch (all infants received comfort measures including swaddling, non-nutritive sucking or were held by parent). Gentle touch was not applied prior to the experimental stimuli. The gentle touch stimulus was provided by a brush stimulator (SENSELab ™ Brush-05, Somedic) applied at a rate of approximately 3 cm/s for 10 seconds prior to heel lancing, across approximately 10 cm of the infant’s lower leg ipsilateral to the heel receiving the lance. Current evidence suggests that this rate (3 cm/s) optimally activates C-tactile (CT) fibres involved in the detection of pleasant touch (Essick et al., 2010; Löken et al., 2009; Triscoli et al., 2014) and our previous study demonstrated the efficacy of gentle touch to reduce brain derived measures following a clinically-required heel lance (Gursul et al., 2018). The experimenter was cued to apply the brushing velocity and noxious stimuli by following a computer visualisation coded using PsychoPy. There was an inter-stimulus interval of approximately one second between the end of the brush stimulation and the heel lance.

The magnitude of the noxious-evoked brain activity and the magnitude of the reflex withdrawal were obtained for each individual trial. Each individual infants’ nociceptive sensitivity was calculated as the mean response to the experimental noxious stimulus. EEG responses were rejected for gross movement artefacts. Following removal of infants whose lances recording were rejected (n=5), 118 out of 492 responses to experimental noxious stimuli were rejected from the EEG analysis. Individuals with seven or less responses to the experimental noxious stimuli were not included in the analysis (n=4) as accurate estimates of baseline nociceptive sensitivity could not be obtained. This left a total of 31 infants (Control Group: n=15, Intervention Group: n=16) for the analysis. Similarly, EMG traces with noise and movement artefacts in the baseline period before the stimulus were rejected. Following removal of infants whose lances recording were rejected (n=7), 18 out of 459 responses to experimental noxious stimuli were rejected, leaving a total of 33 infants (Control Group: n=18, Intervention Group: n=15) included in the EMG analysis. The individual baseline reflex sensitivity was calculated as the median reflex response to the experimental noxious stimulus.

### Study 4: A pharmacological analgesic study

Premature-born infants aged 33 to 43 weeks PMA and due to receive immunisations as inpatients in the neonatal unit were recruited for this study. Infants received Diphtheria, Tetanus, acellular Pertussis, Polio, *Haemophilus influenzae* type b, Hepatitis B (DTaP/IPV/Hib/HepB), Meningococcal group B (MenB) and Pneumococcal (PCV) intramuscular immunisations at eight weeks postnatal age, DTaP/IPV/Hib/HepB immunisations at 12 weeks postnatal age or DTaP/IPV/Hib/HepB, MenB and PCV at 16 weeks postnatal age. Thus, each infant received one or three injections into one or both thighs. For the week 8 and 16 immunisations where MenB vaccine was due, oral paracetamol (15 mg/kg) was administered for the management of pyrexia, in line with the National Institute for Health and Care Excellence (NICE) and British National Formulary for Children (BNFc) guidelines for infants born at less than 4 kg.

A total of 17 infants (Control Group, Figure 5A) were recruited to the study before the clinical practice guidelines were updated in our local neonatal unit to administer paracetamol one-hour prior to the MenB vaccine. Following the guideline change, 16 infants were recruited (Intervention Group), and a subset of 11 of the 16 infants in the Intervention Group also received experimental noxious stimuli before and approximately one-hour after paracetamol was administered (Figure 5A). The average time between paracetamol administration and the second set of experimental noxious stimuli in infants in the Intervention Group subset was 60 minutes (range: 52–70 minutes). The average time between paracetamol administration and immunisation in infants in the Intervention Group was 79 minutes (range: 65–117 minutes). Comfort techniques including swaddling or non-nutritive sucking were used during the immunisation procedures for infants in both groups.

EEG was recorded continuously for the duration of the clinical procedure (i.e immunisation). Needle insertion was time-locked to the EEG recordings using a high-speed video camera (220 frames per second; Firefly MV, Point Grey Research Inc.) that was linked to the recording at the time of acquisition. The time of each individual stimulus was identified retrospectively from the video recordings as the first point of contact of the needle with the skin (Verriotis et al., 2015). Due to technical difficulties (high speed camera failure during set up) and accidental deletion of a recording, three infants were removed from the analysis. Recordings with poor video footage (for which the first point of contact of the needle with the skin was unidentifiable) were rejected from the analysis as well as traces with noise or movement artefact. A total of 29 infants (Control Group: 15 infants, 32 immunisations; Intervention Group: 14 infants, 27 immunisations) were included in the final analysis, with nine infants (18 immunisations) included in the Intervention Group subset. The magnitude of the noxious-evoked brain activity was identified in each individual trial, and for infants in the subset of the Intervention Group baseline nociceptive sensitivity was calculated as the infant’s mean response to the experimental noxious stimuli prior to paracetamol administration. This was compared to the infant’s mean response to the immunisations and mean response to the experimental noxious stimulus recorded one-hour after paracetamol administration.

## Stimulation techniques / clinical procedures

### Experimental noxious stimuli

Experimental noxious stimuli (PinPrick™, MRC Systems) of 64 mN intensity (Study 1) and 128 mN intensity (Studies 3 and 4) were applied to the plantar surface of the heel. The PinPrick™ applies a constant force that stimulates A*∂* and C fibre peripheral nociceptors (Magerl et al., 2001), without piercing the skin. When applied to infants at these forces the stimulus does not cause behavioural distress or clinical concern (Goksan et al., 2015; Hartley et al., 2017, 2015). Stimuli were applied in trains of 10 – 20 trials, with a minimum inter-stimulus interval of 10 seconds (the inter-stimulus interval was increased if necessary, to allow the infant to settle). The 64 mN pinprick stimuli were time-locked to the EEG and EMG recordings using a high-speed video camera (220 frames per second; Firefly MV, Point Grey Research Inc.) that was linked to the recordings at the time of acquisition. The time of each individual stimulus was identified retrospectively from the video recordings with a manual marker when the pinprick’s barrel was first depressed (Hartley et al., 2014; Verriotis et al., 2015). The 128 mN pinprick was automatically time-locked to the EEG and EMG recordings using a contact trigger device (MRC Systems).

### Heel lance

Heel lances were applied only when necessary for blood sampling as part of the infant’s clinical care. Heel lances were automatically time-locked to the EEG and EMG recordings using an event detection interface (Worley et al., 2012). Comfort techniques including swaddling or non-nutritive sucking were used during the heel lance procedures.

## Recording techniques and data preparation

Electrophysiological activity was recorded from DC to 400 Hz using a SynAmps RT 64-channel headbox and amplifiers (Compumedics Neuroscan). CURRYscan7 neuroimaging suite (Compumedics Neuroscan) was used to record activity, with a sampling rate of 2000 Hz. EEG was recorded from eight locations on the scalp (Cz, CPz, C3, C4, Oz, FCz, T3, T4), with reference at Fz and ground at Fpz (forehead) according to the modified international 10-20 system. Preparation gel (Nuprep gel, D.O. Weaver and Co.) was used to gently clean the scalp with a cotton bud before disposable Ag/AgCl cup electrodes (Ambu Neuroline) were placed with conductive paste (Elefix EEG paste, Nihon Kohden). In Study 3, surface electromyography (EMG) was recorded from the limb ipsilateral to the site of stimulation. Bipolar EMG electrodes (Ambu Neuroline 700 solid gel surface electrodes) were placed on the bicep femoris muscle.

EEG signals were filtered from 0.5 – 30Hz with a notch filter at 50 Hz. Epochs were extracted 500 ms before the stimulus and 1000 ms after and were baseline corrected to the pre-stimulus mean. Epochs were rejected if they contained gross movement artefact. Noxious-evoked brain activity was analysed at the Cz electrode for all trials (as this is the electrode site at which the maximal evoked response is observed (Hartley et al., 2017)). The previously validated template of noxious-evoked brain activity was projected onto each individual trial in the time window of interest (400–700 ms after stimulation when the stimulus was applied to the foot, 300–600 ms after stimulation when the stimulus was applied to the thigh – see Supplementary Material) providing a weight indicating the magnitude of the noxious-evoked brain activity (Hartley et al., 2017). Each individual trial was first Woody filtered in the time window of interest to achieve maximum correlation with the template, accounting for individual differences in the latency to the response. A maximum jitter of ± 50 ms for the experimental noxious stimuli and ± 100 ms for the heel lance and immunisations was used for the Woody filtering. In Study 4, additional variation in the latency of the response occurred from the use of the high-speed video camera. To account for this, responses were first Woody filtered within an infant to achieve maximum correlation with the within-subject average.

EMG signals were filtered 10–500 Hz, with a notch filter at 50 Hz and harmonics, and rectified. Epochs were extracted from 2s before to 4s after the stimulus. Individual epochs were rejected due to movement artefact in the baseline period. The data was split into 250 ms windows and the root mean square (RMS) of the reflex signal was calculated in each window. The average RMS across the first four windows after the stimulus (first second after stimulation) was calculated as the magnitude of the reflex withdrawal.

### Statistical analyses

Statistical analysis was performed using MATLAB_R2020a (MathWorks). Linear associations were assessed using Pearson correlation tests in Studies 1, 3 and 4. Statistical significance (alpha<0.05) was assessed non-parametrically via permutation testing with 10,000 permutations using the PALM (Permutation Analysis of Linear Models) toolbox (Winkler et al., 2014). Group differences in Studies 2, 3, and 4 were assessed using linear regressions (unpaired two-sample t-tests, except for the differences in responses to the experimental noxious stimuli before and after paracetamol administration where a paired sample t test was used). When using the paradigm to adjust for the baseline nociceptive sensitivity we used the following linear regression model: *Ŷ* = *b*_0_ + *b*_1_*X*_1_ + *b*_2_*X*_2_, where Ŷ is the magnitude of the response to the clinical procedure, *b*_0_ is the intercept, *X*_1_ is the intervention, and *X*_2_ is the baseline nociceptive sensitivity. Without accounting for baseline nociceptive sensitivity, the model used was *Ŷ* = *b*_0_ + *b*_1_*X*_1_ . Statistical significance (alpha<0.05) in Studies 3 and 4 group comparisons was assessed non-parametrically via permutation testing with 10,000 permutations using PALM. The difference between the intervention and control group in Study 4 was assessed using a linear mixed effects model, with subject and number of immunisations set as random effects. Two-sided tests were used for all statistical analysis with a significance level of 0.05.

## Author contributions

MC, DG, CH, FM and RS conceived the study; MC, DG, CH, FA, MvdV, GSM, MB, REF, GG, AH and FM acquired the data; MC, CH and DG analysed the data; MC, CH, DG, LB, ED, RR, FM and RS interpreted the data. MC, CH, FM and RS wrote the manuscript. GSM, FA, REF, AH, GG, EA and FM had clinical oversight of the studies. CH, FM and RS had oversight and leadership responsibility. All authors critically reviewed the manuscript.

## Acknowledgments

We would like to thank the infants and their parents for taking part in this study. CH is a Wellcome Trust/Royal Society Sir Henry Dale Fellow (213486/Z/18/Z).

## Data availability

The data that support the findings of this study, including data to produce figures 1, 3, 4, 5 and supplementary figures 1 to 5, are available from the corresponding author upon reasonable request. Data sharing requests should be directed to.

## Code availability

The magnitude of noxious-evoked brain activity in response to the experimental noxious stimuli and clinically-required procedures was calculated using the template of noxious evoked brain activity previously validated for experimental and clinical stimuli and available from (Hartley et al., 2017).

