## Supplementary Materials for "Quantifying individual nociceptive sensitivity to optimise analgesic trials in infants"

### Supplementary Materials and Methods

#### Validating the template of noxious-evoked brain activity – accounting for latency differences when the stimuli is applied to different body sites

The template of noxious-evoked brain activity has previously been validated for experimental and clinical stimuli applied to the heel (Hartley et al., 2017). As other clinical interventions like immunisations are injected into the infant's thigh, the latency to the brain activity response is expected to be shorter compared with stimuli applied to the foot. In an independent sample of 17 infants aged 36 to 42 weeks' PMA (demographic details given in Table 1), we investigated the latency of the noxious-evoked brain activity following experimental noxious stimuli applied to the foot, thigh and hand. A total of 10-12 experimental noxious stimulus (128 mN, inter-stimulus interval of at least 10s) were applied to the infant's hand (n = 17 infants), foot (n = 17) and thigh (n = 10). The order of the stimulus location and the side were randomly selected by the research team before each test occasion (right = 8, left = 9). In 7 studies, the EEG recordings were linked to a high-speed camera (Firefly MV, Point Grey Research Inc.) to time-lock the experimental stimuli (Hartley et al., 2015). In the other 10 studies, the stimuli were time-locked to the EEG recordings using a contact trigger device (MRC systems).

The EEG signal was filtered 0.5-30 Hz with a notch filter at 50 Hz. Epochs were extracted 500 ms before the stimulus and 1000 ms after (total 1500 ms per epoch), and the traces were baseline corrected to the pre-stimulus mean. Noxious-evoked brain activity was analysed at the Cz electrode for all the trials. Individual EEG epochs with artefacts were removed from the analysis and infants with less than 5 trials on any individual locations were removed from the analysis for that location. The final foot, thigh, and hand analysis included 14, 8 and 14 infants respectively.

Data from all individual trials from all infants were Woody filtered in the 0–700 ms interval after the stimulus onset with a maximum shift of +/-50ms (aligning to the average of the data). The average response for each subject to the stimulus and the average background were calculated from the Woody filtered data. Clusters of timepoints where the noxious stimulus was significantly different from background were identified using a nonparametric cluster analysis (Maris and Oostenveld, 2007). The cluster-based test statistic was calculated from 1000 random permutations of the data, and the threshold for cluster significance was set as the 97.5 percentile of the permuted data. The midpoint of the cluster was identified and the time window for the Principal Component Analysis (PCA) was taken as the 300 ms window about this midpoint, rounding to the nearest 100 ms, for each of the responses to stimuli applied to the hand, foot and thigh separately. PCA was performed to identify the Principal Components (PCs) of the activity in response to the stimuli compared with the background activity (Bromm and Scharein, 1982; Fabrizi et al., 2011; Hartley et al., 2016; Slater et al., 2010b) and the PC weights compared between background and the stimulus response using a paired t-test. The first two PCs accounted for over 73% of the variance in the data across all stimulus conditions and were the only components tested.

The PC in which the weights were significantly different in response to the noxious stimulus compared with background activity was selected as the noxious-evoked response. This PC and the previously described template of noxious-evoked brain activity (Hartley et al., 2017) were compared using correlation, to demonstrate the validity of using the template to identify noxious-evoked brain activity for stimuli applied to different body locations.

Consistent with previous studies (Hartley et al., 2017, 2015), experimental noxious stimuli applied to the foot evoked a cluster of activity that was significantly different from background activity in the time window from 456 – 654 ms after the stimulus ($p = 0.014$, cluster-corrected nonparametric test, Figure S3A). Principal component analysis (PCA) applied in the time window 400 – 700 ms identified a representative waveform of the noxious-evoked response - the second principal component (PC) had significantly higher weights following the noxious stimulation compared to background brain activity ($p = 0.035$, Figure S3B) and was significantly correlated with the validated template of noxious-evoked brain activity ($r = 0.97$, $p < 0.001$, Figure S3C). Experimental noxious stimuli applied to the hand evoked a cluster of activity that was significantly different from background activity in the time window 298 – 461 ms ($p = 0.01$, Figure S3D) (Kasser et al., 2019). PCA was applied in the time window 200 – 500 ms following stimulation; the weights of the second PC were significantly higher following the experimental noxious stimuli compared with background activity ($p < 0.001$, Figure S3E) and this PC was highly correlated with the template of noxious-evoked brain activity ($r = 0.97$, $p < 0.001$, Figure S3F).

Experimental noxious stimuli applied to the thigh evoked a cluster of activity that was significantly different from background activity in the time window 280 – 563 ms ($p= 0.002$, Figure S3G). PCA was applied in the time window 300 – 600 ms following stimulation; the weights of the second PC were significantly higher following the experimental noxious stimuli compared with background activity ($p < 0.001$, Figure S3H) and this PC was highly correlated with the template of noxious-evoked brain activity ($r = 0.98$, $p < 0.001$, Figure S3I). Overall, the latencies of noxious-evoked brain activity are related to the physical distance of the stimulus location from the brain (Figure S3J) as expected. With immunisation applied to the thigh the latency to the response is expected to be approximately 300 ms.

**Validating the template - for use in studies of immunisation**

To validate the suitability of the template (in the time-window from 300 – 600 ms) to characterise immunisation-evoked brain activity we compared the activity evoked by the immunisation with the background brain activity in the Control Group in Study 4 ($n = 15$) who did not receive paracetamol prior to immunisation. Clusters of timepoints where the noxious stimulus was significantly different from background were identified using a nonparametric cluster analysis, with 1000 random permutations of the data, to check that significant noxious-evoked activity was observed in the same time window as that observed in response to experimental noxious stimuli applied to the thigh. Immunisation evoked activity was significantly different to background in the time window 416 – 594 ms ($p = 0.035$, non-parametric cluster analysis, Figure S4A) following stimulation. PCA in the time window 300 – 600 ms identified the first PC weights (which accounted for 58% of the variance) as significantly higher following immunisation compared with background activity ($p = 0.004$, Figure S4B) and this PC was highly correlated with the template of noxious-evoked brain activity ($r = 0.97$, $p < 0.001$, Figure S4C). An event-related potential with a similar waveform and latency has been previously recorded in one and two month old term-born infants following immunisations (Verriotis et al., 2015). Therefore, the template of noxious-evoked brain activity, derived in an independent sample of infants (Hartley et al., 2017), was considered appropriate to characterise response to immunisation and used in the subsequent analysis.

### Supplementary Figures

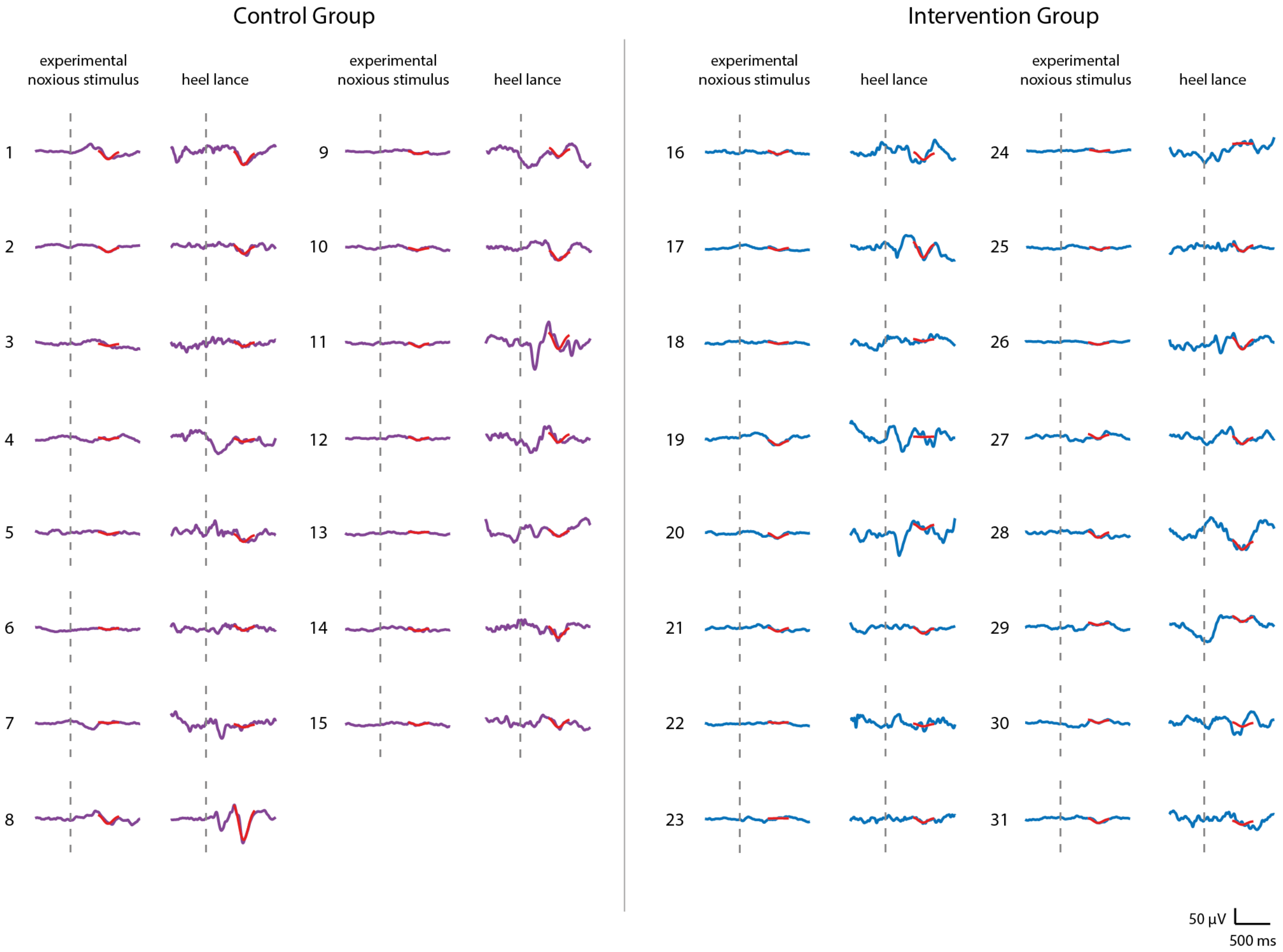


**Fig. S1. Noxious-evoked brain activity in individual infants in the Control Group and Intervention Group, Study 3.** Average noxious-evoked brain activity to the experimental noxious stimuli and the corresponding brain activity following heel lancing within the 31 infants in Study 3. The template of noxious-evoked brain activity (Hartley et al., 2017) is overlaid in red and the grey dashed lines indicate the point of stimulation. Infants in the intervention group (EEG traces in blue) received gentle brushing before the heel lancing.

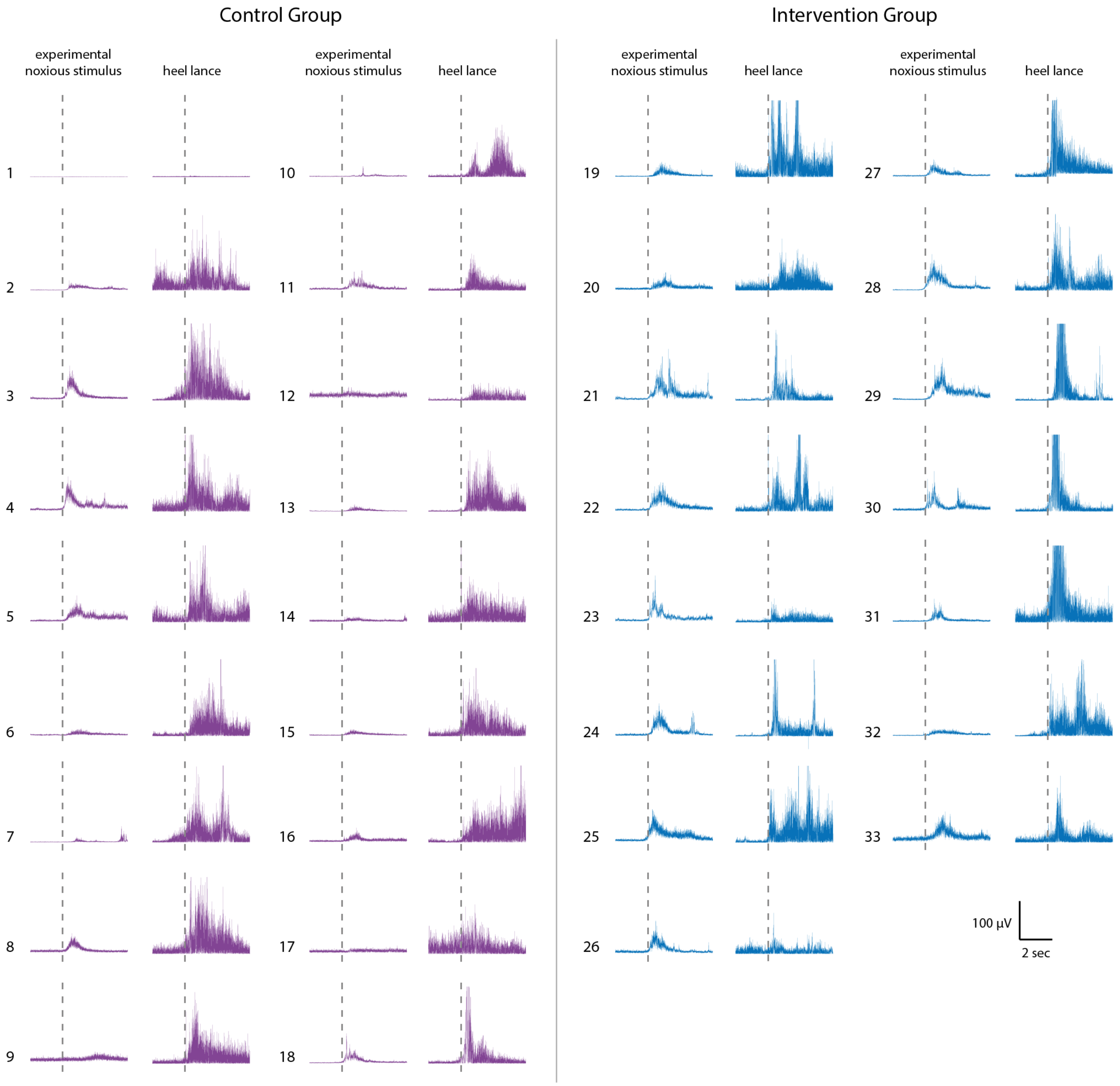


**Fig. S2. Reflex withdrawal activity in individual infants in the Control Group and Intervention Group, Study 3.** Average reflex withdrawal in response to experimental noxious stimulation and reflex withdrawal evoked by heel lancing in the 31 infants included in Study 3. Infants in the intervention group (EMG traces in blue) received gentle brushing before the heel lancing.

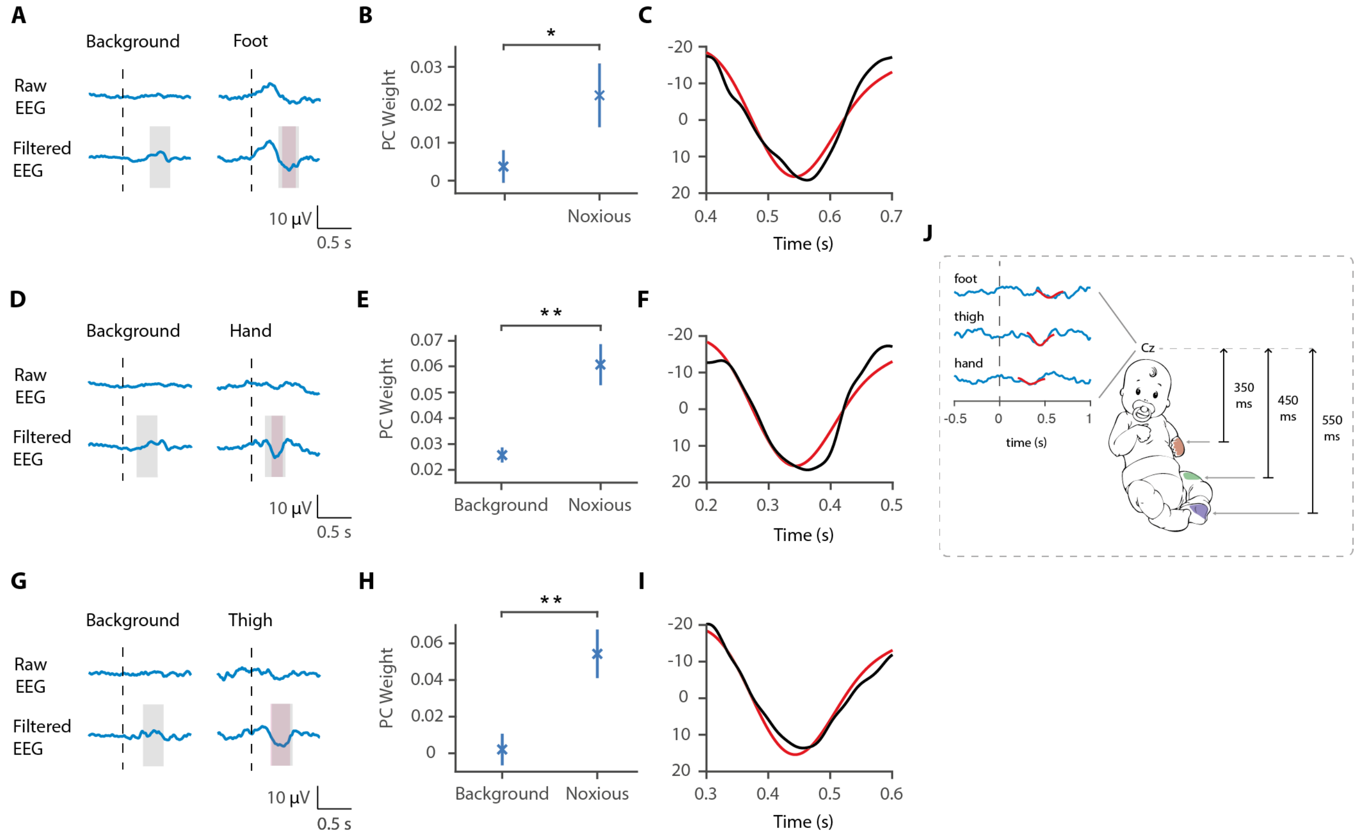


**Fig. S3**. **Latency of the noxious-evoked brain activity in response to stimulation on the foot, thigh and hand. (A)** Raw average and Woody filtered EEG following experimental noxious stimuli of the foot and during background brain activity. **(B)** Principal Component Analysis was conducted in the time window 400 – 700 ms after the stimulus and the second principal component weights were significantly higher to noxious stimulation of the foot compared with in the background brain activity (paired t-test, * $p<0.05$). **(C)** The second principal component waveform (black) was highly correlated with the previously described template of noxious-evoked brain activity (red). **(D)** Raw average and Woody filtered EEG following experimental noxious stimuli of the hand and during background brain activity. **(E)** Principal Component Analysis was conducted in the time window 200 – 500 ms after the stimulus and the second principal component weights were significantly higher to noxious stimulation of the hand compared with in the background brain activity (** $p < 0.001$). **(F)** The second principal component waveform (black) was highly correlated with the previously described template of noxious-evoked brain activity (red). **(G)** Raw average and Woody filtered EEG following experimental noxious stimuli of the thigh and during background brain activity. **(H)** Principal Component Analysis was conducted in the time window 300 – 600 ms after the stimulus and the second principal component weights were significantly higher to noxious stimulation of the thigh compared with in the background brain activity (** $p<0.001$). **(I)** The second principal component waveform (black) was highly correlated with the previously described template of noxious-evoked brain activity (red). **(J)** Schematic representation of the latency of the noxious-evoked brain activity with stimuli applied across different locations. In panels (A,D,G) dashed lines show the time point of the stimulation. Pink shading shows the time window of the identified noxious response cluster and grey shading shows the time window where the PCA was performed.

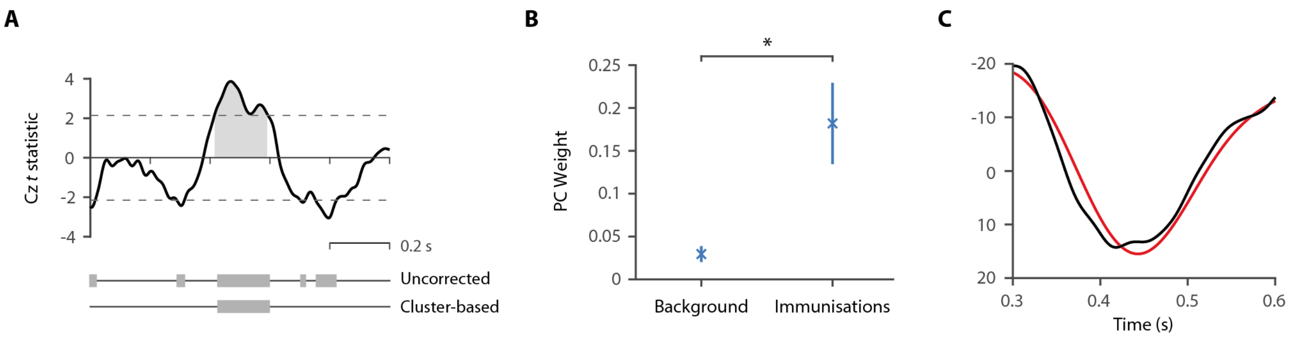


**Fig. S4**. **Validation of the template for use in immunisation studies. (A)** *t* statistics from the comparison of the noxious-evoked brain activity following immunisations and during background brain activity in 15 infants from the Control Group in Study 4. Dashed lines indicate the t statistic threshold for cluster significance, set as the 97.5 percentile of the permuted data. The grey bars indicate time periods outside of the t statistic threshold and the significant time window identified with the cluster analysis is illustrated by the grey shading area. **(B)** Principal Component Analysis was conducted in the time window 300 – 600 ms after the immunisations and the first principal component weights were significantly higher following immunisations compared with in the background brain activity (* $p < 0.01$). **(C)** The first principal component waveform (black) was highly correlated with the previously described template of noxious-evoked brain activity (red).

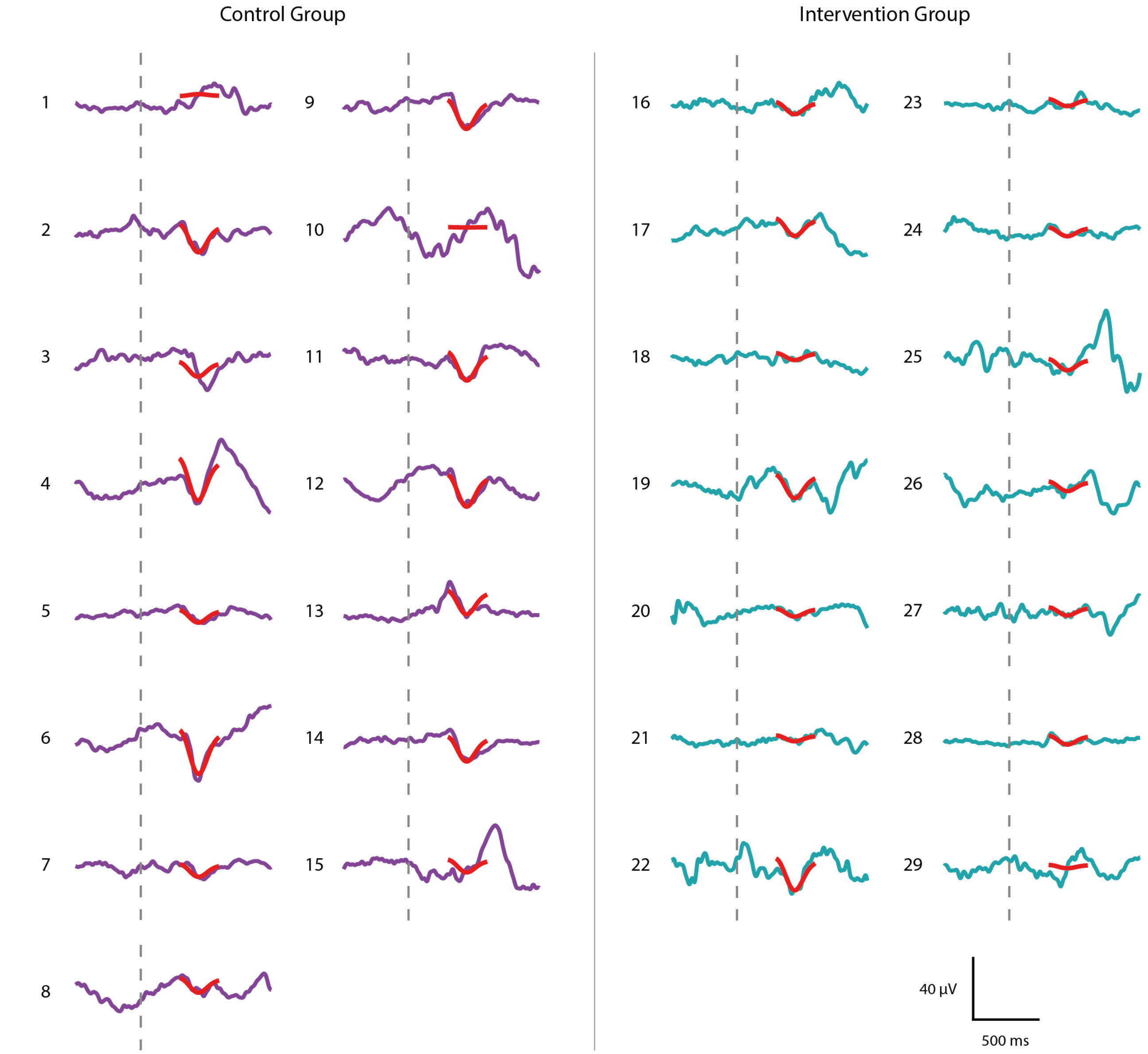


**Fig. S5. Noxious-evoked brain activity following immunisations in individual infants in the Control Group and Intervention Group, Study 4.** Average noxious-evoked brain activity in individual infants following immunisations in the 29 infants in Study 4. The template of noxious-evoked brain activity (Hartley et al., 2017) is overlaid in red and the grey dashed lines indicate the point of stimulation. Infants in the Intervention Group (EEG traces in teal) received paracetamol approximately one-hour prior to the immunisations.
